# COVID-19 Related Mortality and The BCG Vaccine

**DOI:** 10.1101/2020.05.01.20087411

**Authors:** Jan A. Paredes, Valeria Garduño, Julian Torres

## Abstract

The coronavirus disease 2019 (COVID-19) pandemic has become a worldwide emergency. In the attempt to search for interventions that would improve outcomes, some researchers have looked at the potential benefit of BCG vaccination. These early studies have found a statistically significant reduction in COVID-19 related mortality in countries with a current universal bacille Calmette-Guérin (BCG) vaccination policy; partially explained by induced heterologous immunity. However, just as the authors themselves noted, the nature of ecological studies make them very prone to the presence of several confounders. This paper tries to answer the question as to whether a statistically significant difference in mortality rates exists between countries with differing BCG vaccination policies; while being the first to try to account for most of these confounders. We compared the number of COVID-19 related deaths per 1 million inhabitants as well as the number of deaths at the time the countries hit the 1000^th^ COVID-19 case. Countries were divided in those which never had a BCG vaccination policy, those with a prior vaccination policy and those with a current vaccination policy. All data was gathered from publicly available sources. It was found that no statistically significant difference exists in mortality rates between countries with differing BCG vaccination policies. This result seems to reflect the notion that heterologous immunity fades with time after administration. Nevertheless, the immunostimulatory potential of the BCG vaccine might still prove useful in the development of future vaccines or other prophylactic measures.

## Introduction

The coronavirus disease 2019 (COVID-19) is the result of infection by the novel coronavirus SARS-CoV-2; previously 2019-nCoV. First detected in late December 2019 in Wuhan, China; it has since spread to 6 continents and is estimated to have infected over 2.1 million people and caused over 145,000 deaths worldwide as of April 16, 2020.^1, 2, 3^ The rapid spread of the disease around the world has become a worldwide emergency. The high mortality of the disease in certain populations (i.e. the elderly), along with the lack of vaccinations or effective antiviral treatment have resulted in significant efforts in trying to find interventions that would improve outcomes.^4, 5^ Early studies have shown an inverse relationship between the bacille Calmette-Guérin (BCG) vaccination policy of countries and COVID-19 related mortality. Countries with a current universal BCG vaccination policy seem to have significantly lower mortality rates when compared to countries that do not implement BCG in their childhood vaccination schedules.^6, 7, 8^ The immunostimulant properties of the vaccine could in theory improve outcomes through heterologous immunity, which refers to the cross-protection the immune system confers between two unrelated pathogens. ^9^

Past studies support the notion of BCG induced heterologous immunity as they have found a BCG-associated reduction in all cause mortality within the first years of life.^10,11^ This reduction in mortality has been found to be more noticeable in regions with high childhood mortality and infectious disease burden such as West Africa.^12,13^ Similarly, the rates of infection by non mycobacterial pathogens, were found to be lower in children vaccinated with BCG.^14^ In a study conducted by Hollm et al. in 2014, more than 150,000 children in 33 different countries were found to have a risk reduction of 17% to 37% for the development of acute lower respiratory infections.^15^

All these findings seem to suggest that the BCG might indeed be associated to improved outcomes of COVID-19. However, some studies debate the efficacy of the heterologous immunity induced by the BCG vaccine. The Danish Calmette Study, conducted by Lone Graff et al., found no differences in the rates of infection or hospitalization in children up to the age of 15 months; partially explained by the lower infection disease burden in the region.^16^ Moreover, the protective effect conferred by the BCG seems to wane with time and a previous study conducted by Yu et al. in 2007 found no cross protection between BCG and the Severe Acute Respiratory Syndrome Coronavirus (SARS-CoV), which belongs to the same family as SARS-CoV-2.^14, 17^ Regarding the negative association between universal BCG vaccination and COVID-19 related mortality, several confounders were identified by the authors themselves. Under-reporting and reduced SARS-CoV-2 capability testing in low income countries, along with comparing mortality rates at different times in each countries’ epidemiological curve, could all serve as potential confounders.^6,7,8^

This paper tries to account for these potential confounders for the first time. This study would then serve in answering the question as to whether a statistically significant difference in mortality rates exists between countries with differing BCG vaccination policies. We gathered data from publicly available sources and compared the number of COVID-19 related deaths per 1 million inhabitants as well as the number of deaths at the time the countries hit the 1000^th^ COVID-19 case. To avoid any possible bias on each country’s capability for testing, reporting and their resources available for healthcare, only high income countries were compared.^18^ Our findings suggest no statistical difference exists between countries with different vaccination programs.

## Material and Methods

The study conducted is an observational ecological study, comparing the mortality rates of countries with differing BCG vaccination programs. The data retrieved included: total population, total number of COVID-19 cases, total number of COVID-19 attributable deaths, COVID-19 attributable deaths per 1 million population, COVID-19 attributable deaths at the 1000^th^ case, percentage of the population over 65 years old, and the year of introduction and cessation of the BCG vaccine where applicable.

The number of total COVID-19 cases, total COVID-19 attributable deaths, deaths per 1 million population and deaths at the 1000^th^ case were retrieved from the Worldometer website.^3^ Data on the total population, percentage of the population over 65 years old and the GDP income was retrieved from The World Bank Data Website.^19^ Data on BCG vaccination policies was retrieved from The BCG World Atlas.^20^ The IBM SPSS program was then used to analyze statistical data.

The inclusion criteria were countries with:

- At least 1,000 COVID-19 cases per the Worldometer website.
- At least 1 million inhabitants per The World Bank Data Website.
- A Gross National Income (GNI) per capita higher than 12,375 US dollars, per The World Bank Data Website.

Countries with no data available were excluded from the study.

Countries that met all criteria were divided in three groups:

1. Countries with no history of universal BCG vaccination policy.
2. Countries with an actual vaccination policy.
3. Countries with a prior universal BCG vaccination policy.

For the third group, countries that had a universal BCG vaccination policy introduced in 1955 or earlier that lasted until at least 1975 were included (as to ensure that the elderly population would have been vaccinated).

For each group, the mean and standard deviation of COVID-19 attributable deaths per 1 million population and number of deaths at the 1000^th^ case were calculated from the combined data of each country in the respective group.

First compared was mean and standard deviation of the number of COVID-19 attributable deaths per 1 million population. Countries with no history of universal vaccination programs were compared to the combined data of countries with a current or prior vaccination policy using the *t-*test. Then, the three groups were compared separately using the ANOVA test. Afterwards, the number of COVID-19 attributable deaths at the 1000^th^ was compared following the same method, presuming a normal distribution data where pertinent.

Finally, adjustments were made to account for differences in demographics. The mean and standard deviation was calculated for the percentage of the population over 65 years old for all the countries included in the study. The previous comparisons were repeated, this time excluding countries that strayed more than 1.5 standard deviations from the mean.

## Results

After analysis, 20 countries met the inclusion criteria for the study. Out of these, 5 high income countries never had a universal BCG vaccination policy (table 1), 5 currently have a universal BCG vaccination policy and 10 had a previous BCG vaccination policy introduced in 1955 or earlier; with its respective withdrawal no earlier than 1975 (table 2).

**Table 1:** Regions with No Prior Universal BCG Vaccination Policy. Data on BCG vaccination policies was retrieved from The BCG World Atlas. Total population data was retrieved from The World Bank Data. The number of COVID–19 cases and COVID–19 attributable mortality were retrieved from the Worldometer website.

| Country | Population<br>(in<br>thousands) | Total Cases | Total<br>Deaths | Deaths/1M | Deaths at<br>1000th case | % >65<br>years old |
| --- | --- | --- | --- | --- | --- | --- |
| Canada | 37,057 | 30,106 | 1,195 | 32 | 12 | 17.2 |
| USA | 326,687 | 677,570 | 34,617 | 105 | 30 | 16 |
| Italy | 60,421 | 168,941 | 22,170 | 367 | 29 | 22.8 |
| Netherlands | 17,231 | 29,214 | 3,315 | 193 | 20 | 18.9 |
| Belgium | 11,433 | 34,809 | 4,857 | 419 | 10 | 18.7 |
| **Total** | **452,829** | **940,640** | **66,154** | **1116** | **101** |  |

**Table 2.** Regions with Current or Prior Universal BCG Vaccination Policy. Data on BCG vaccination policies was retrieved from The BCG World Atlas. Total population data was retrieved from The World Bank Data. The number of COVID–19 cases and COVID–19 attributable mortality were retrieved from the Worldometer website.

| **Country** | **Population (in thousands)** | **Total Cases** | **Total Deaths** | **Deaths/1 M Pop** | **Deaths at 1000th case** | **% >65 years old** | **Year of BCG introduction** | **Year of BCG cessation** |
| --- | --- | --- | --- | --- | --- | --- | --- | --- |
| Israel | 8,882 | 12,758 | 142 | 16 | 1 | 11.3 | 1955 | 1982 |
| Finland | 5,515 | 3,369 | 75 | 14 | 7 | 21.9 | 1941 | 2006 |
| Sweden | 10,175 | 12,540 | 1,333 | 132 | 3 | 19.9 | 1940 | 1975 |
| Denmark | 5,793 | 6,879 | 321 | 55 | 4 | 19.6 | 1946 | 1986 |
| Czech Republic | 10,629 | 6,433 | 169 | 16 | 0 | 19.6 | 1953 | 2010 |
| Austria | 8,840 | 14,476 | 410 | 46 | 3 | 18.8 | 1952 | 1990 |
| Slovenia | 2,073 | 1,268 | 61 | 29 | 28 | 19.6 | 1947 | 2005 |
| France | 66,977 | 165,027 | 17,920 | 275 | 16 | 20.3 | 1950 | 2007 |
| UK | 66,460 | 103,093 | 13,729 | 202 | 21 | 18.3 | 1953 | 2005 |
| Australia | 24,982 | 6,468 | 63 | 2 | 7 | 15.8 | 1950's | 1980's |
| Croatia | 4,087 | 1,791 | 35 | 9 | 7 | 20.4 | 1948 | Current |
| Hungary | 9,775 | 1,652 | 142 | 15 | 66 | 19.3 | 1953 | Current |
| Japan | 126,529 | 9,231 | 190 | 2 | 35 | 28.2 | 1942 | Current |
| Poland | 37,974 | 7,918 | 314 | 8 | 14 | 17.5 | 1955 | Current |
| Chile | 18,729 | 8,807 | 105 | 5 | 24 | 11.8 | 1948 | Current |

The mean number of deaths per 1 million population for countries with no universal BCG vaccination (223.2 ± 166.1) was not statistically significant from countries with current or previous BCG vaccination programs (55 ± 82.5) [95% Confidence Intervals (CI) (−34.8, 371); p = 0.85]. When comparing the means of the 3 groups separately, a statistically significant difference was noted between countries with a current BCG vaccination program (7.8 ± 4.8) and countries that never implemented a universal BCG vaccination policy (223.2 ± 166.1) [95% CI (−386.5, −44.2); p = .013].

The mean number of deaths at the 1000th case for countries with current or previous BCG vaccination programs and countries that never implemented BCG vaccination in their schedules was 15.7 ± 17.5 and 20.2 ± 9.2 respectively. No statistically significant difference was noted between these two groups [95% CI (−12.9, 21.8); p value = .597]. When comparing the mean number of deaths of the three different groups, a statistically significant difference was noted between the means of countries with an actual BCG program (29.2 ± 23.1) and countries with previous BCG programs (9 ± 9.4) [95% CI (0.66, 39.7); p value = .042]. (Graph 1)

**Graph 1.**
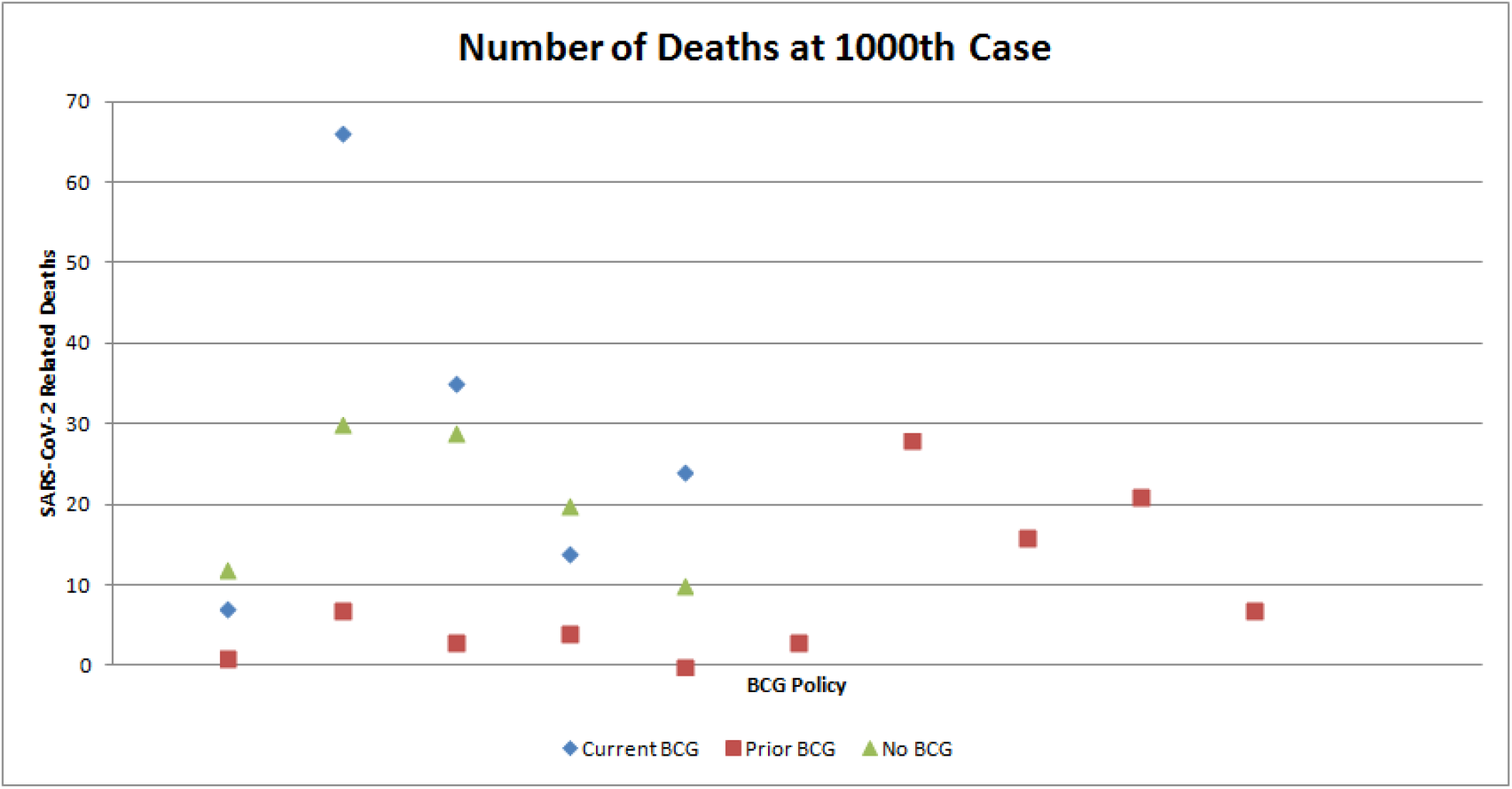
Number of SARS-CoV-2 Related Deaths at the 1000th Case. Data on BCG vaccination policies was retrieved from The BCG World Atlas. The number of COVID-19 cases and COVID-19 attributable mortality were retrieved from the Worldometer website. No statistically significant difference was found when comparing mortality at the 1000th case. (p = > 0.05).

It is important to note that some countries have a different constitution in the age of their population. The mean percentage and standard deviation of adults aged >65 in the 20 countries was 18.7 ± 3.6; Japan (28.2%), Israel (11.3%) and Chile (11.8%) were more than 1.5 SD’s away from the mean. When comparing the number of deaths at the 1000th case and the total number of deaths per 1 million population of the three groups, no statistically significant difference was found when these countries were excluded from the analysis (p = > 0.05).

## Discussion

The results found in this study seem to suggest no statistically significant difference in mortality arises from BCG vaccination. This is in contrast to ealy studies that found an inverse relationship between mortality rates and BCG vaccination status.^6,7,8^ A striking difference in deaths per 1 million population was noted between current BCG using countries and countries that never implemented a universal vaccination program, which is similar to the results found in previous studies. However, just as the WHO noted, this doesn’t reflect demographic differences, disease burden and the different stages of the pandemic in these countries.^21^

When trying to account for confounders by comparing the number of deaths at a particular point in time and considering demographic differences, no statistically significant difference was found. Furthermore, the number of deaths at the 1000th case were actually lower in countries that had a prior BCG vaccination program (9 ± 9.4) when compared to countries with an actual universal vaccination policy (29.2 ± 23.1). Nevertheless, this finding was brought into question as no difference was found when accounting for demographic differences in these countries.

These results seem to fit in line with previous findings regarding BCG-induced heterologous immunity. Findings seem to suggest an increase in immune response, however this is highly dependent on the time the research subjects received the BCG vaccine. The population that has been previously shown to benefit from BCG-induced heterologous immunity is restricted to the pediatric population.^10-15, 22-23^ Furthermore, this benefit seems to only be apparent when the regions involved already have high childhood mortality, as evidenced by the Danish Calmette Study.^16^ The pediatric population can contract COVID-19 and even serve as potential carriers because of their mostly asymptomatic course.^24, 25^ However, this fact alone is not enough evidence to suggest a change in the vaccination program of countries that do not implement BCG in their schedules. This if further emphasized by the apparent lack of cross protection the BCG vaccine confers against coronaviruses. This was evidenced by a study conducted by Yu et al. in 2007 when he looked at the potential protection childhood vaccines conferred against SARS-CoV.^17^

Although childhood vaccination seems to have no effect on mortality rates of the adult population, the BCG vaccine could still prove useful. The early genome sequencing of SARS-CoV-2 is helping current efforts in developing a vaccine against it. The use of recombinant technology could help with the early development of a vaccine by using vectors and embedding them with SARS-CoV-2 antigens.^26^ Recombinant BGC vaccines could help in this field as they induce a considerable immune response and have been used before as vectors to non-mycobacterial related antigens.^27-29^ Also of interest is the potential effect revaccination might have on the elderly population. The potential of BCG-induced heterologous immunity after revaccination has shown promise, as demonstrated by Warhana et al.^30^ Two current clinical trials are looking into this by vaccinating healthcare workers with BCG to see if there is any potential benefit.^31, 32^

There are some limitations to be considered in this study. Although great care was taken to try to avoid as many confounders as possible, some still remain. The difference in mortality seems to be higher in the male population than in the female population, which could affect the overall mortality in countries with differing gender compositions.^33^ Also, some populations are more genetically susceptible to certain diseases and complications than others (e.g. Latin Americans and diabetes).^34^ Some researchers are looking into this by analyzing genetic differences in the expression of the ACE2 receptor or the CCL2 and MBL genes, which could affect susceptibility to SARS-CoV-2.^6, 35, 36^ Finally, each country’s response to the outbreak affects subsequent policies taken by other countries and although the coverage of countries currently using BCG was over 90% no data was available for countries with prior vaccination programs.^20^

## Conclusion

The status of BCG vaccination policy does not seem to have an effect in COVID-19 related mortality. Although data is difficult to analyze due to the presence of multiple confounders, statistically significant differences seem to disappear once much of the confounders are taken into consideration. Nevertheless, the immunostimulant properties of the BCG vaccine could prove useful in future developments of a vaccine using recombinant technology. Likewise, revaccination with BCG has shown promise in the past for the induction of heterologous immunity. Current medical trials are underway looking at the potential benefits of vaccinating healthcare workers.

## Data Availability

All relevant data are within the manuscript and its Supporting Information files.

http://datahelpdesk.worldbank.org/knowledgebase/articles/906519-world-bank-country-and-lending-groups

http://www.bcgatlas.org

https://www.worldometers.info/coronavirus/coronavirus-cases/

## Conflicts of interest

No conflicts of interest to disclose.

## Notes

### Competing Interest Statement

The authors have declared no competing interest.

### Funding Statement

The authors received no specific funding for this work.

### Author Declarations

All relevant ethical guidelines have been followed

## References

1. Xiaolu Tang, Changcheng Wu, Xiang Li, Yuhe Song, Xinmin Yao, Xinkai Wu, Yuange Duan, Hong Zhang, Yirong Wang, Zhaohui Qian, Jie Cui, Jian Lu, On the origin and continuing evolution of SARS-CoV-2, National Science Review, nwaa036, https://doi.org/10.1093/nsr/nwaa036

2. Di Wu, Tiantian Wu, Qun Liu, Zhicong Yang. The SARS-CoV-2 outbreak: what we know. International Journal of Infectious Diseases, 2020; DOI: 10.1016/j.ijid.2020.03.004

3. Coronavirus Cases. Worldometer website. Last updated april 16, 2020. Accessed: April 16, 2020. Available at: https://www.worldometers.info/coronavirus/coronavirus-cases/

4. Jason Oke, Carl Heneghan. Global Covid-19 Case Fatality Rates. Centre for Evidence Based Medicine, Published March 17, 2020. Last Updated April 7, 2020. Accessed April 10, 2020. Available at:https://www.cebm.net/covid-19/global-covid-19-case-fatality-rates/

5. Weilong Shang, Yi Yang, Yifan Rao, Xiancai Rao. The outbreak of SARS-CoV-2 pneumonia calls for viral vaccines. npj Vaccines; 2020 (5(1):1–3”)20200310.

6. Anita Shet, Debashree Ray, Neelika Malavige, Mathuram Santosham, Naor Bar-Zeev. Differential COVID-19-attributable mortality and BCG vaccine use in countries. doi: 10.1101/2020.04.01.20049478

7. Aaron Miller, Mac Josh Reandelar, Kimberly Fasciglione, Violeta Roumenova, Yan Li, Gonzalo H Otazu. Correlation between universal BCG vaccination policy and reduced morbidity and mortality for COVID-19: an epidemiological study. March 28, 2020. doi: 10.1101/2020.03.24.20042937

8. Gursel M, Gursel I. Is Global BCG Vaccination Coverage Relevant To The Progression Of SARS-CoV-2 Pandemic? [published online ahead of print, 2020 Apr 6]. Med Hypotheses. 2020;109707. doi:10.1016/j.mehy.2020.109707

9. Kleinnijenhuis J, Kleinnijenhuis J, Quintin J, Preijers F, Benn C. S, Joosten L. A. B, Jacobs C, van Loenhout J, Xavier R. J, Aaby P, van der Meer J. W. M, van Crevel R, Netea M. G: Long-Lasting Effects of BCG Vaccination on Both Heterologous Th1/Th17 Responses and Innate Trained Immunity. J Innate Immun 2014;6:152–158. doi: 10.1159/000355628

10. Evidence based recommendations on non-specific effects of BCG, DTP-containing and measles-containing vaccines on mortality in children under 5 years of age. World Health Organization website. https://www.who.int/immunization/sage/meetings/2014/april/1_NSE_Backgroundpaper_final.pdf?ua=1,. Published 2014; Accessed April 9, 2020.

11. Butkeviciute E, Jones CE, Smith SG. Heterologous effects of infant BCG vaccination: potential mechanisms of immunity. Future Microbiol. 2018;13(10):1193–1208. doi:10.2217/fmb-2018-0026

12. Butkeviciute E, Jones CE, Smith SG. Heterologous effects of infant BCG vaccination: potential mechanisms of immunity. Future Microbiol. 2018;13(10):1193-1208. doi:10.2217/fmb-2018-0026

13. Vaugelade J, Pinchinat S, Guiella G, Elguero E, Simondon F: Non-specific effects of vaccination on child survival: prospective cohort study in Burkina Faso. BMJ 2004;329:1309.

14. De Castro MJ, Pardo-Seco J, Martinón-Torres F. Nonspecific protection of neonatal BCG vaccination against hospitalization due to respiratory infection and sepsis. Clin. Infect. Dis. 2015;60(11):1611–1619.

15. Hollm-Delgado M-G, Stuart EA, Black RE. Acute lower respiratory infection among Bacille Calmette-Guérin–vaccinated children. Pediatrics. 2014;133(1):e73–e81

16. Lone Graff Stensballe, Henrik Ravn, Nina Marie Birk, Jesper Kjærgaard, Thomas Nørrelykke Nissen, Gitte Thybo Pihl, Lisbeth Marianne Thøstesen, Gorm Greisen, Dorthe Lisbeth Jeppesen, Poul-Erik Kofoed, Ole Pryds, Signe Sørup, Peter Aaby, Christine Stabell Benn, BCG Vaccination at Birth and Rate of Hospitalization for Infection Until 15 Months of Age in Danish Children: A Randomized Clinical Multicenter Trial, Journal of the Pediatric Infectious Diseases Society, Volume 8, Issue 3, July 2019, Pages 213–220, https://doi.org/10.1093/jpids/piy029.

17. Yu Y, Jin H, Chen Z, et al. Children’s vaccines do not induce cross reactivity against SARS-CoV. J Clin Pathol. 2007;60(2):208–211. doi:10.1136/jcp.2006.038893

18. New country classifications by income level: 2019–2020. World Bank Blogs website: https://blogs.worldbank.org/opendata/new-country-classifications-income-level-2019-2020 Published July 01, 2019. Accessed April 16, 2020.

19. World Bank Country and Lending Groups, Country Classification. The World Bank Data website: http://datahelpdesk.worldbank.org/knowledgebase/articles/906519-world-bank-country-and-lending-groups. Accessed April 16, 2020.

20. The BCG World Atlas, 2nd Edition. Available at: http://www.bcgatlas.org/ Updated 2017, Accessed April 16, 2020.

21. Bacille Calmette-Guérin (BCG) vaccination and COVID-19, Scientific Brief. World Health Organization, April 12, 2020. Accessed on April 18, 2020.

22. Higgins JP, Soares-Weiser K, López-López JA, et al. Association of BCG, DTP, and measles containing vaccines with childhood mortality: systematic review [published correction appears in BMJ. 2017 Mar 8;356:j1241]. BMJ. 2016;355:i5170. Published 2016 Oct 13. doi:10.1136/bmj.i5170

23. Garly ML, Martins CL, Bale C, Balde MA, Hedegaard KL, Gustafson P, Lisse IM, Whittle HC, Aaby P: BCG scar and positive tuberculin reaction associated with reduced child mortality in West Africa: a non-specific beneficial effect of BCG? Vaccine 2003;21:2782–2790.

24. Qiu H, Wu J, Liang H, Yunling L, Song Q, Chen D. Clinical and epidemiological features of 36 children with coronavirus disease 2019 (COVID-19) in Zhejiang, China: an observational cohort study. Lancet Infect Dis. 2020; published online March 25, 2020. Available at: https://doi.org/10.1016/S1473-3099(20)30198-5

25. Alyson A Kelvin, Scott Halperin. COVID-19 in children: the link in the transmission chain. The Lancet Infectious Diseases, published:March 25, 2020; DOI:https://doi.org/10.1016/S1473-3099(20)30236-X

26. Richard Lane. Sarah Gilbert: carving a path towards a COVID-19 vaccine. The Lancet, Volume 395, Issue 10232, P1247, April 18, 2020. DOI: 10.1016/S0140-6736(20)30796-0

27. Fennelly GJ, Flynn JL, Meulen V, Liebert UG, Bloom BR. Recombinant bacille calmette-guréin priming against measles. J Infect Dis. (1995) 172:698–705. 10.1093/infdis/172.3.698

28. Wang H, Liu Q, Liu K, Zhong W, Gao S, Jiang L, et al. Immune response induced by recombinant *Mycobacterium bovis* BCG expressing ROP2 gene of *Toxoplasma gondii*. Parasitol Int. (2007) 56:263–8. 10.1016/j.parint.2007.04.003

29. Palavecino CE, Cespedes PF, Gomez RS, Kalergis AM, Bueno SM. Immunization with a recombinant bacillus Calmette-Guerin strain confers protective Th1 immunity against the human metapneumovirus. J Immunol. 2014;192(1):214–223.

30. Wardhana Datau EA, Sultana A, Mandang VV, Jim E. The efficacy of Bacillus Calmette-Guerin vaccinations for the prevention of acute upper respiratory tract infection in the elderly. Acta Med Indones. 2011 Jul;43(3):185–90.

31. Reducing Health Care Workers Absenteeism in Covid-19 Pandemic Through BCG Vaccine (BCG-CORONA). https://clinicaltrials.gov/ct2/show/NCT04328441.

32. BCG Vaccination to Protect Healthcare Workers Against COVID-19 (BRACE). https://clinicaltrials.gov/ct2/show/NCT04327206.

33. Clare Wenham Julia Smith Rosemary Morgan. COVID-19: the gendered impacts of the outbreak. The Lancet, Volume 395, Issue 10227, P846–848, March 14, 2020 DOI: 10.1016/S0140-6736(20)30526-2

34. Hispanic/Latino Americans and Type 2 Diabetes. Centers for Disease Control And Prevention website: https://cdc.gov/diabetes/library/features/hispanic-diabetes.html. Accessed April 18, 2020

35. Tu X, Chong WP, Zhai Y, et al. Functional polymorphisms of the CCL2 and MBL genes cumulatively increase susceptibility to severe acute respiratory syndrome coronavirus infection. The Journal of infection. 2015;71(1):101–109.

36. Cao Y, Li L, Feng Z, et al. Comparative genetic analysis of the novel coronavirus (2019-nCoV/SARS-CoV-2) receptor ACE2 in different populations. Cell Discov. 2020;6:11.

